# SARS-CoV-2 RNA screening in routine pathology specimens

**DOI:** 10.1101/2021.01.25.21250082

**Authors:** Saskia von Stillfried, Sophia Villwock, Roman D. Bülow, Sonja Djudjaj, Eva M. Buhl, Angela Maurer, Nadina Ortiz-Brüchle, Peter Celec, Barbara M. Klinkhammer, Dickson W.L. Wong, Claudio Cacchi, Till Braunschweig, Ruth Knüchel-Clarke, Edgar Dahl, Peter Boor

## Abstract

Virus detection methods are important to cope with the SARS-CoV-2 pandemics. Apart from the lung, SARS-CoV-2 was detected in multiple organs in severe cases. Less is known on organ tropism in patients developing mild or no symptoms, and some of such patients might be missed in symptom-indicated swab testing.

Here we tested and validated several approaches and selected the most reliable RT-PCR protocol for the detection of SARS-CoV-2 RNA in patients’ routine diagnostic formalin-fixed and paraffin-embedded (FFPE) specimens available in pathology, to assess a) organ tropism in samples from COVID-19-positive patients, b) unrecognized cases in selected tissues from negative or not-tested patients during a pandemic peak, and c) retrospectively, pre-pandemic lung samples.

We identified SARS-CoV-2 RNA in four samples from confirmed COVID-19 patients, in two gastric biopsies, one colon resection, and one pleural effusion specimen, while all other specimens, particularly from patients with mild COVID-19 disease course, were negative. In the pandemic peak cohort, we identified one previously unrecognized COVID-19 case in tonsillectomy samples. All pre-pandemic lung samples were negative.

In conclusion, SARS-CoV-2 RNA detection in FFPE pathology specimens can potentially improve surveillance of COVID-19, allow retrospective studies, and advance our understanding of SARS-CoV-2 organ tropism and effects.

## Introduction

Identification and isolation of infected individuals with severe acute respiratory syndrome coronavirus-2 (SARS-CoV-2) is an effective preventive measure to limit the 2019 coronavirus disease (COVID-19) pandemic. Previous studies suggested that between 40-45% of patients infected with SARS-CoV-2 develop only mild symptoms or even remain asymptomatic (Oran and Topol, 2020). Some patients may initially present with very mild respiratory or atypical, e.g., gastrointestinal, symptoms (Wang, et al., 2020). This could limit the effectiveness in identifying infected individuals if the examination is indicated only by the presence of symptoms. Apart from the nasal and respiratory tract and the lung (Hou, et al., 2020), several organs have been described as positive for viral RNA, especially the salivary gland, heart, liver, central nervous system, kidneys, lymph nodes, spleen, and colon (Azzi, et al., 2020, Lamers, et al., 2020, Puelles, et al., 2020, Sekulic, et al., 2020).

Pathologists analyze a wide variety of samples from virtually all tissues, most of which are formalin-fixed and paraffin-embedded (FFPE). The latter is an efficient method for long-term preservation of proteins and nucleic acids but also inactivates infectious agents, including SARS-CoV-2. Therefore, the analysis of FFPE specimens does not require the high biosafety precautions required for swabs or unfixed specimens. A smaller proportion of specimens analyzed in pathology, particularly for perioperative diagnostics, biobanking, and some cytologic analyses, are processed unfixed and are thus potentially infectious (Guerini-Rocco, et al., 2020). To date, the majority of SARS-CoV-2 RNA analyses have been performed on FFPE autopsy specimens or cell pellets (Liu, et al., 2020, Puelles, et al., 2020). For the detection of SARS-CoV-2 in non-autopsied tissues, mainly case reports on a small number of individual patients and very few studies with more than 20 cases focusing on single organs have been published so far (Escher, et al., 2020, Smithgall, et al., 2020). In this single-center study, we established a SARS-CoV-2 RNA detection protocol for FFPE material to a) assess the feasibility of detecting viral RNA in samples from clinically diagnosed COVID-19 patients, b) evaluate the potential use of FFPE samples to screen for previously unrecognized infected patients, and c) consider the use of archival material, e.g. to screen for potential cases before identification of the first local index patients, as recently proposed (Deslandes, et al., 2020). To search for previously unrecognized infected patients, given the previous data on the traceability of SARS-CoV-2 RNA in various tissues and organs, we focused on samples from oropharyngeal and sinonasal mucosa, salivary glands, lung, colon, and kidney (Borczuk, et al., 2020, Guerini-Rocco, et al., 2020, Puelles, et al., 2020, Remmelink, et al., 2020, Sekulic, et al., 2020). One of the first and most affected SARS-CoV-2 hotspots in Germany was in the catchment area of our center in Aachen, with 22% asymptomatic patients (Streeck, et al., 2020). Therefore, our Institute of Pathology was well-positioned to address the above research questions using FFPE material from our diagnostic archive.

## Results

### Evaluation of various RT-PCR SARS-CoV-2 RNA detection systems for FFPE specimens screening

Using a SARS-CoV-2 RNA standard with a defined viral copy number (2-fold dilution), we validated two different one-step RT-PCR methods (RealStar and TaqMan) in single- and multiplex approaches with different primer and probe sets for SARS-CoV-2 RNA detection (Figure 1a), and evaluated RNA detection in FFPE samples in this context. The RealStar method is a simple single-tube assay that detects the SARS-CoV-2 S gene and the B-βCoV E gene in a multiplex approach as a dual-target assay. We measured amplification and, using linear regression, found that RT-PCR efficiency for the SARS-CoV-2 S gene was within the range of efficient RT-PCR (90-110%) (Taylor, et al., 2010). In contrast, the efficiency of the B-βCoV E gene was outside this range (<90%). Using the TaqMan method, we analyzed the RT-PCR efficiencies of the SARS-CoV-2 E gene, the RdRp gene, and the N gene as previously suggested (Corman, et al., 2020), in a singleplex approach for selecting two assays to establish a dual-target assay. We found that the E- and RdRp-gene assays were within the range of efficient RT-PCR, but the N-gene assay was out of this range (>110%). Therefore, we additionally analyzed the RT-PCR efficiency of the E- and RdRp-gene in a multiplex approach as a potential method to save resources, but we found that the efficiency decreased in the multiplex approach (Figure 1b). In addition to RT-PCR efficiency, we determined the theoretical limit of detection by the y-axis intercept of the linear regression, i.e., the Ct value of the smallest detectable unit of a viral copy number μL^-1^. The detection limit of the RealStar multiplex and TaqMan singleplex approach was similar for all assays, but for the TaqMan multiplex approach, the detection limit decreased compared to the singleplex approach (Figure 1c). In general, we observed higher Ct values (2-5 Ct values) for detection of the RdRp gene in the SARS-CoV-2 RNA standard dilution compared to detection of the E gene at the same viral copy number. Based on the Ct values of the SARS-CoV-2 RNA standard dilution series, we additionally found that the multiplex RealStar approach became inaccurate at <100 viral copy numbers μL^-1^. With the singleplex TaqMan approach, detection of SARS-CoV-2 RNA by the E and RdRp genes was accurate up to 3 viral copy numbers μL^-1^, whereas detection by the N gene became inaccurate at <25 copies μL^-1^. The multiplex TaqMan approach with E and RdRp gene became inaccurate at <100 viral copy numbers μL^-1^ (Table 1). To evaluate RT-PCR methods in FFPE samples, we used diluted RNA (1:50) isolated from the trachea and lung of clinically confirmed COVID-19 autopsy cases. We tested the RealStar multiplex and TaqMan singleplex approaches for the E-gene assay and found that in the same samples, SARS-CoV-2 RNA detection was negative with the RealStar method but positive with the TaqMan method (Table 2).

**Table 1:**
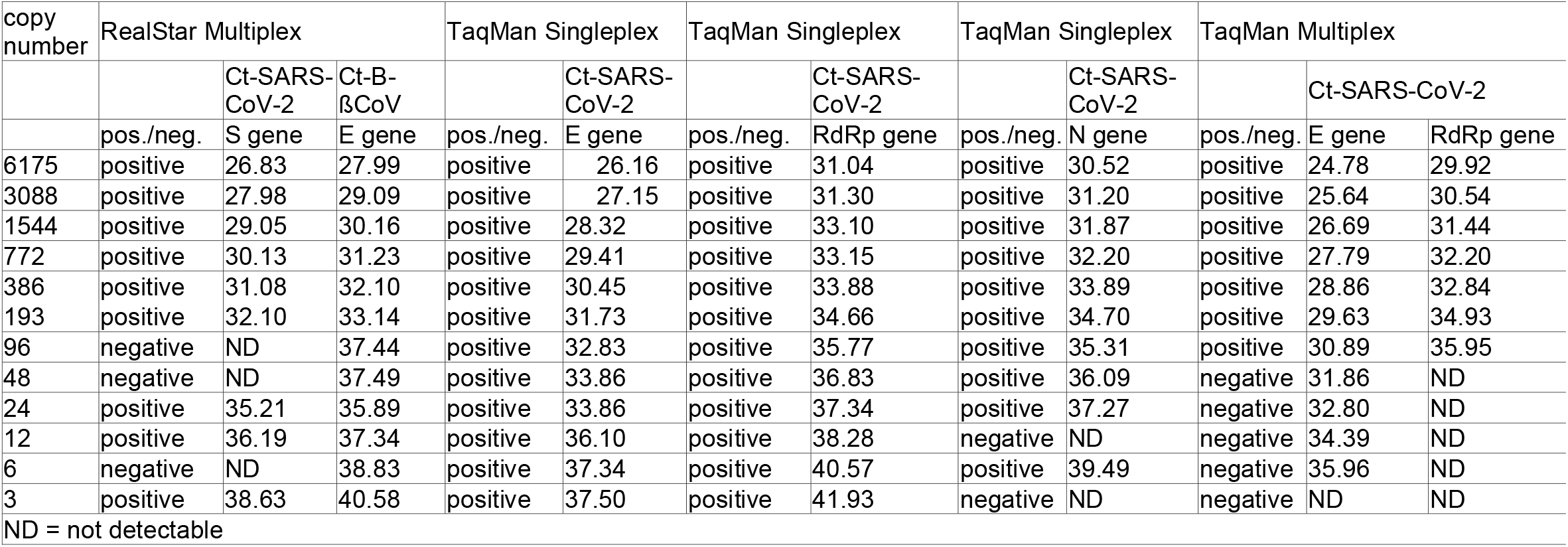
Detection of SARS-CoV-2 in RNA standard dilution series by the various RT-PCR methods. Ct values for SARS-CoV-2 RNA detection using RealStar multiplex, TaqMan singleplex (E, RdRp, and N gene), and TaqMan multiplex (E, RdRp gene) assays in 2-fold SARS-CoV-2 RNA standard dilutions with defined copy numbers. Only TaqMan E and RdRp gene assays are accurate to 3 μL^-1^ viral copy numbers.

| copy number | RealStar Multiplex |  |  | TaqMan Singleplex |  | TaqMan Singleplex |  | TaqMan Singleplex |  | TaqMan Multiplex |  |  |
| --- | --- | --- | --- | --- | --- | --- | --- | --- | --- | --- | --- | --- |
| | | Ct-SARS-CoV-2 | Ct-B-βCoV | | Ct-SARS-CoV-2 | | Ct-SARS-CoV-2 | | Ct-SARS-CoV-2 | | Ct-SARS-CoV-2 | |
|  | pos./neg. | S gene | E gene | pos./neg. | E gene | pos./neg. | RdRp gene | pos./neg. | N gene | pos./neg. | E gene | RdRp gene |
| 6175 | positive | 26.83 | 27.99 | positive | 26.16 | positive | 31.04 | positive | 30.52 | positive | 24.78 | 29.92 |
| 3088 | positive | 27.98 | 29.09 | positive | 27.15 | positive | 31.30 | positive | 31.20 | positive | 25.64 | 30.54 |
| 1544 | positive | 29.05 | 30.16 | positive | 28.32 | positive | 33.10 | positive | 31.87 | positive | 26.69 | 31.44 |
| 772 | positive | 30.13 | 31.23 | positive | 29.41 | positive | 33.15 | positive | 32.20 | positive | 27.79 | 32.20 |
| 386 | positive | 31.08 | 32.10 | positive | 30.45 | positive | 33.88 | positive | 33.89 | positive | 28.86 | 32.84 |
| 193 | positive | 32.10 | 33.14 | positive | 31.73 | positive | 34.66 | positive | 34.70 | positive | 29.63 | 34.93 |
| 96 | negative | ND | 37.44 | positive | 32.83 | positive | 35.77 | positive | 35.31 | positive | 30.89 | 35.95 |
| 48 | negative | ND | 37.49 | positive | 33.86 | positive | 36.83 | positive | 36.09 | negative | 31.86 | ND |
| 24 | positive | 35.21 | 35.89 | positive | 33.86 | positive | 37.34 | positive | 37.27 | negative | 32.80 | ND |
| 12 | positive | 36.19 | 37.34 | positive | 36.10 | positive | 38.28 | negative | ND | negative | 34.39 | ND |
| 6 | negative | ND | 38.83 | positive | 37.34 | positive | 40.57 | positive | 39.49 | negative | 35.96 | ND |
| 3 | positive | 38.63 | 40.58 | positive | 37.50 | positive | 41.93 | negative | ND | negative | ND | ND |

ND = not detectable

**Table 2:** SARS-CoV-2 RNA detection in FFPE tissues. Isolated and diluted (1:50) RNA from FFPE tissue (trachea, lung) from clinically confirmed COVID-19 autopsy cases are negative for SARS-CoV-2 in the RealStar multiplex approach but positive at 20-30 viral copy numbers μL^-1^ in the TaqMan singleplex (E gene) approach.

| Tissue | dilution | RealStar Multiplex |  |  | TaqMan Singleplex |  |  |
| --- | --- | --- | --- | --- | --- | --- | --- |
| | | pos./neg. | Ct-SARS-CoV-2<br>S gene | Ct-B-βCoV<br>E gene | pos./neg. | Ct-SARS-CoV-2<br>E gene | viral copy<br>number $\mu\text{L}^{-1}$ |
| Trachea | 1:50 | negative | ND | ND | positive | 34.50 | 28 |
| Lung | 1:50 | negative | ND | ND | positive | 34.88 | 22 |
| Lung | 1:50 | negative | ND | ND | positive | 34.45 | 29 |

**Figure 1:**
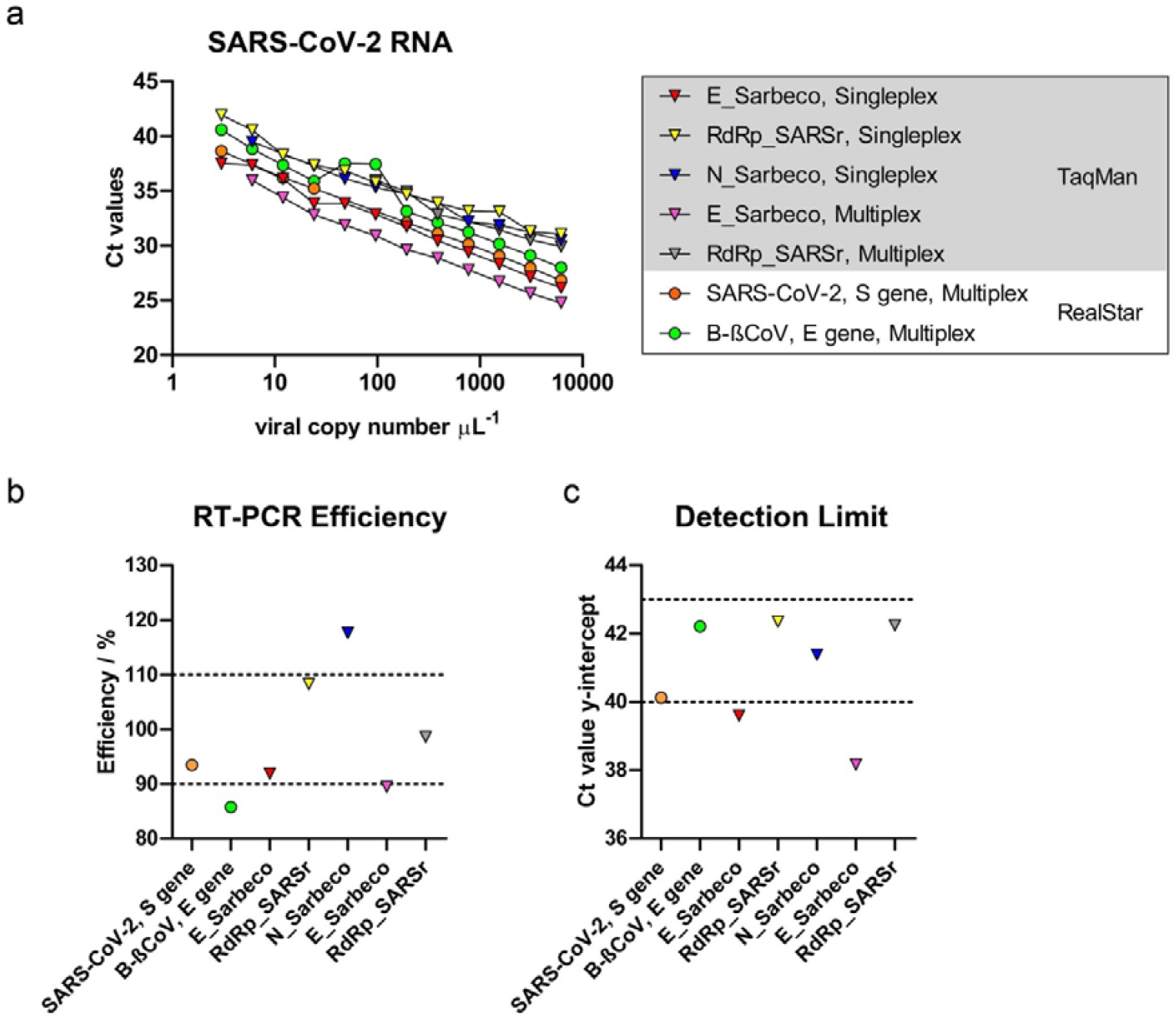
Evaluation of RT-PCR efficiency and detection limit of the different primer and probe sets with the two different RT-PCR methods by SARS-CoV-2 RNA standard. Ct values of primer and probe sets as single and multiplex approaches using RealStar and TaqMan methods in 2-fold dilution series of SARS-CoV-2 RNA standard. b) Using linear regression of each assay, RT-PCR efficiency was calculated according to the equation 100x(−1+10^−1/slope^). For successful RT-PCR assays, the efficiency ranges from 90 to 110% (Taylor, et al., 2010). The limit of detection was determined by the y-axis intercept of the linear regression, where the smallest unit of a copy number is detectable (10^0^ = 1) (Vogels, et al., 2020). The limits of detection vary between a Ct value of 38 and 42, depending on the assay.

Therefore, we established a workflow for SARS-CoV-2 RNA detection using the TaqMan RT-PCR singleplex approach with the E gene assay for screening RNA isolated from FFPE tissue samples from patients. For confirmatory testing (dual-target assay), we established the RdRp gene assay (Supplementary Figure 2).

**Figure 2:**
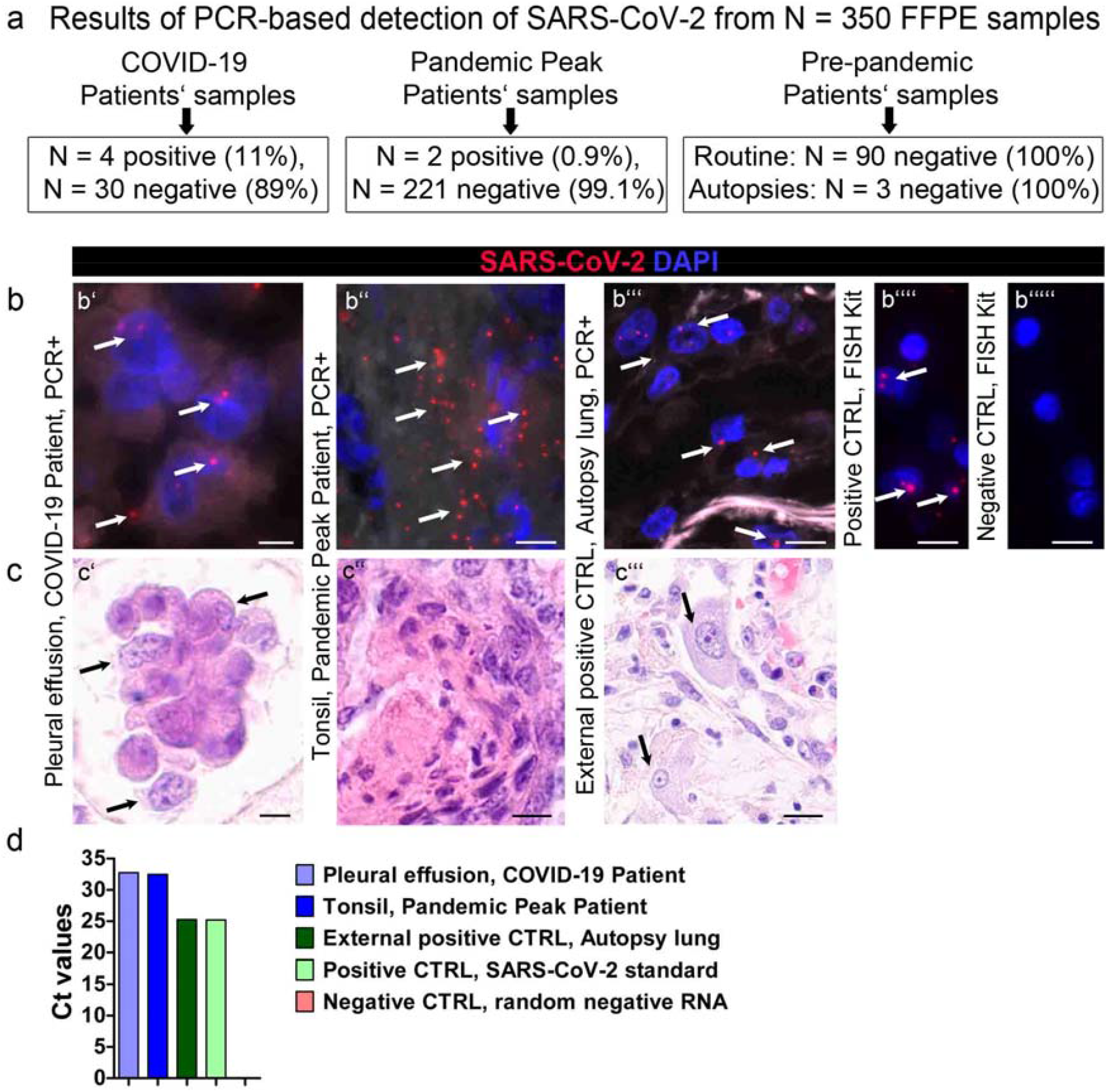
Results of RT-PCR-based detection of SARS-CoV-2 RNA in the three cohorts studied (N = 350) and method validation using fluorescence in situ hybridization (FISH). (a) SARS-CoV-2 RT-PCR detection in all three cohorts resulted in four positive samples in COVID-19 patient samples and one positive sample in pandemic peak patient samples. (b-c) Method validation by FISH and hematoxylin-eosin staining; (b’) SARS-CoV-2 RNA-positive pleural effusion sample (arrows, patient 12, Table 4) and (b’’) SARS-CoV-2-positive tonsil sample from pandemic patient with viral RNA in detritus-filled crypts (arrows); b’’) Lung tissue from an autopsy case with clinically confirmed SARS-CoV-2 infection showed a red fluorescent signal of SARS-CoV-2 RNA (arrows, scale bar = 5 µm), b’’’’) Positive control with a red fluorescent signal of Homo sapiens POLR2A gene (arrows, scale bar = 10 µm), b’’’’’) Negative control (dap gene from Bacillus subtilis, scale bar = 10 µm). c’) Light micrograph of pleural effusion sample showing a group of reactive macrophages (arrow, HE, scale bar = 10 µm). c’’) Light micrograph of tonsil sample in b’’); c’’’) Light micrograph of lung tissue in b’’’) with reactive macrophages (arrows, HE, scale bar = 20 µm). d) RT-PCR results (individual SARS-CoV-2 E-gene Ct values) of the samples.

### Detection of SARS-CoV-2 RNA in the COVID-19 Patients Cohort

In the COVID-19-positive patient cohort, the relatively small number of tissue samples (n = 34 samples from 22 patients) was not surprising because invasive procedures are kept to a minimum in COVID-19 patients. The 34 samples included two gastric biopsy specimens, one colon resection specimen, and one FFPE specimen from a cytospin preparation of a pleural effusion in which SARS-CoV-2 RNA was detectable (Figure 2a, 2d; Table 3). Using fluorescence *in situ* hybridization (FISH)), intracytoplasmic SARS-CoV-2 RNA was detectable in cells morphologically identified as macrophages (Figure 2b’), supported by hematoxylin-eosin staining (Figure 2c’). This sample, as well as autopsy lung samples from COVID-19 patients (Figure 2b’’’, 2c’’’, 2d), allowed us to confirm the specificity of the RT-PCR method with an independent FISH approach using a different target RNA sequence. All remaining samples were negative for SARS-CoV-2 RNA (Tables 3 and 4). Notably, in patients with mild disease progression (i.e. not requiring mechanical ventilation) and early COVID-19 stage, SARS-CoV-2 RNA was not detectable in soft tissues, normal and neoplastic oral mucosa, lymph nodes, salivary gland, ovarian and peritoneal lavage fluid, placenta, and a lung biopsy. In patients with severe disease (i.e. requiring mechanical ventilation) and early COVID-19, samples from soft tissues, thymoma, and also from pleural effusion were negative for SARS-CoV-2 RNA. Morphologically, no characteristic features of viral infection or sequelae of viral infection were evident in extrapulmonary tissues. Lung samples from three patients with onset of COVID-19 symptoms long before surgery, i.e., 50-81 days, showed pulmonary morphologies consistent with the severity of clinical COVID-19 disease progression, e.g. focal fibrosis in a mild disease course (patient 10, Table 3, Figure 3a’ and 3a’’) and diffuse fibrosis with nearly complete obliteration of the alveolar spaces in severe disease courses (patient 17, Table 4, Figure 3b’ and 3b’’). Confounding factors such as previous lung disease, radiation to the lungs, or a history of tobacco abuse were not known to exist in these patients.

**Table 3:** Patient characteristics of COVID-19 Positive Patients with mild disease course. Clinically confirmed COVID-19 patients with mild COVID-19 infection without mechanical ventilation. Time of surgery/sampling was set as zero, times are given as days -/+ = before/after surgery/sampling.

|  | Patient Nr. | Sex | Age range | Duration of ventilation (d) / hospitalization (d) | First symptoms (d +/- sampling) | Positive test for SARS-CoV-2 (d +/- sampling; localization) |  | SARS-CoV-2 RNA from FFPE | Viral copy number/ µl | Localization | Indication for surgery/ sampling | Specimen received as | Histologic result |
| --- | --- | --- | --- | --- | --- | --- | --- | --- | --- | --- | --- | --- | --- |
|  |  |  |  |  |  | First | Last |  |  |  |  |  |  |
| Mild disease course without mechanical ventilation | 1 ‡ | F | 81-85 | 0/27 | asymptomatic | 7; upper airways | 18; upper airways | neg | . | right kidney | neoplasia | formalin-fixed resection | urothelial cancer |
|  | 2 ‡ | M | 61-65 | 0/18 | 3 | 3; upper airways |  | neg | . | left lateral oral floor | neoplasia | perioperative fresh-frozen section | tumor-free oral mucosa |
|  |  |  |  |  |  |  |  | neg | . | anterior oral floor |  | formalin-fixed resection | SCC of oral mucosa |
|  |  |  |  |  |  |  |  | neg | . | left lymph node level IIa |  | perioperative fresh-frozen section | lymph node |
|  |  |  |  |  |  |  |  | neg | . | right lymph node level IIa |  | perioperative fresh-frozen section |  |
|  |  |  |  |  |  |  |  | neg | . | left submandibular gland |  | perioperative fresh-frozen section | salivary gland |
|  | 3 ‡ | F | 31-35 | 0/1 | asymptomatic | 0; upper airways |  | neg | . | right ovary | neoplasia | perioperative fresh-frozen section | borderline tumor of ovary |
|  |  |  |  |  |  |  |  |  | . | left ovary |  | formalin-fixed resection |  |
|  |  |  |  |  |  |  |  |  | . | peritoneal cavity |  | peritoneal lavage fluid |  |
|  | 4 ‡ | M | 71-75 | 0/9 | unknown | 1; upper airways |  | pos | 4 | gastric antrum | gastritis | formalin-fixed biopsy | chronic antrum gastritis, no intestinal metaplasia, no helicobacter pylori |
|  | 5 ‡ | M | 51-55 | 0/18 | asymptomatic | -1; upper airways |  | pos | 922 | gastric antrum | gastritis | formalin-fixed biopsy | chronic antrum gastritis, intestinal metaplasia, no helicobacter pylori |
|  | 6 ‡ | F | 31-35 | 0/2 | asymptomatic | -1; upper airways |  | neg | . | uterine cavity | missed abortion | formalin-fixed resection | placenta and endometrium |
|  | 7 ‡ | F | 66-70 | 0/26 | asymptomatic | -11; upper airways |  | neg | . | left upper thigh soft tissue | impaired wound healing | formalin-fixed resection | soft tissue with low-grade inflammation |
|  | 8 ‡ | M | 76-80 | 0/18 | -21 | -18; upper airways | 4; upper airways | neg | . | upper left lung lobe bronchus mucosa | Mediastinal nodule | formalin-fixed resection | inflammatory bronchial and peribronchial tissue |
|  | 9 ‡ | M | 66-70 | 0/68 | -30 | -29; upper airways |  | neg | . | left and right liver lobe | transplant | formalin-fixed resection | liver cirrhosis |
|  | 10 ‡ | M | 61-65 | 0/5 | -66 | -60; upper & lower airways |  | neg | . | lingular lymph node | neoplasia | perioperative fresh-frozen section | Lymph node MET of RCC |
|  |  |  |  |  |  |  |  | neg | . | left lung lingula |  | perioperative fresh-frozen section | Lung MET of RCC |
|  |  |  |  |  |  |  |  | neg | . | lower left lung lobe |  | formalin-fixed resection | DAD, organizing with acute inflammation |

‡ = dismissed; DAD = diffuse alveolar damage; FFPE = formalin-fixed paraffin-embedded; MET = metastasis; RCC = renal cell carcinoma

**Table 4:**
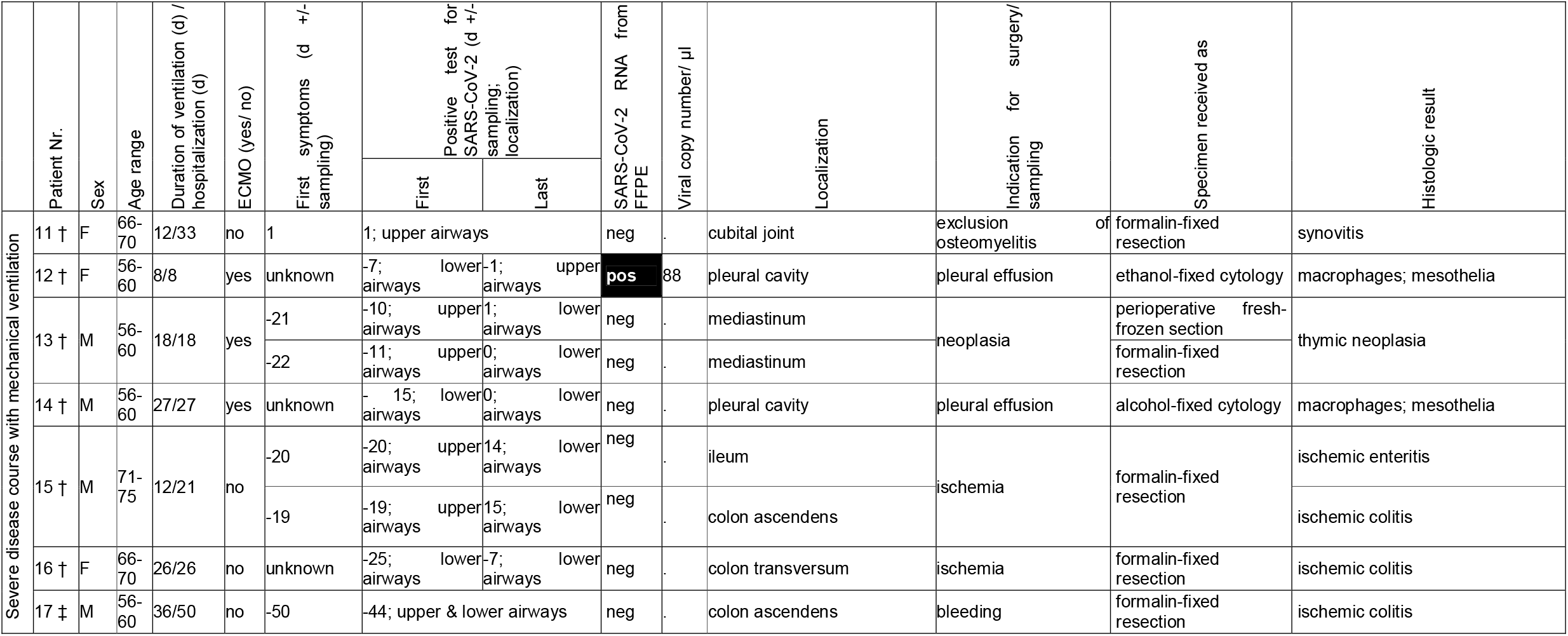
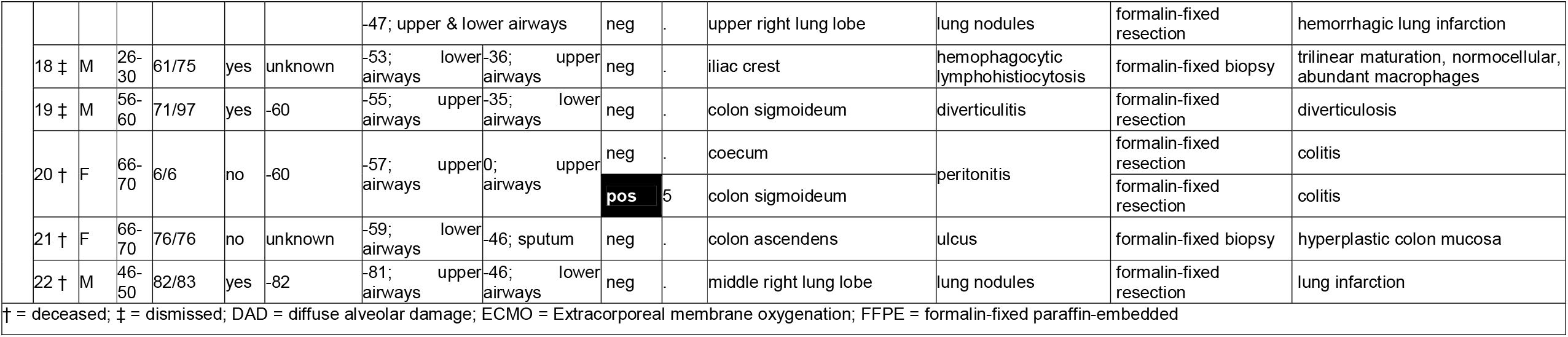
Patient characteristics of COVID-19 Positive Patients with severe disease course. Clinically confirmed COVID-19 patients with severe COVID-19 infection on mechanical ventilation. Time of surgery/sampling was set as zero, times are given as days -/+ = before/after surgery/sampling.

**Figure 3:**
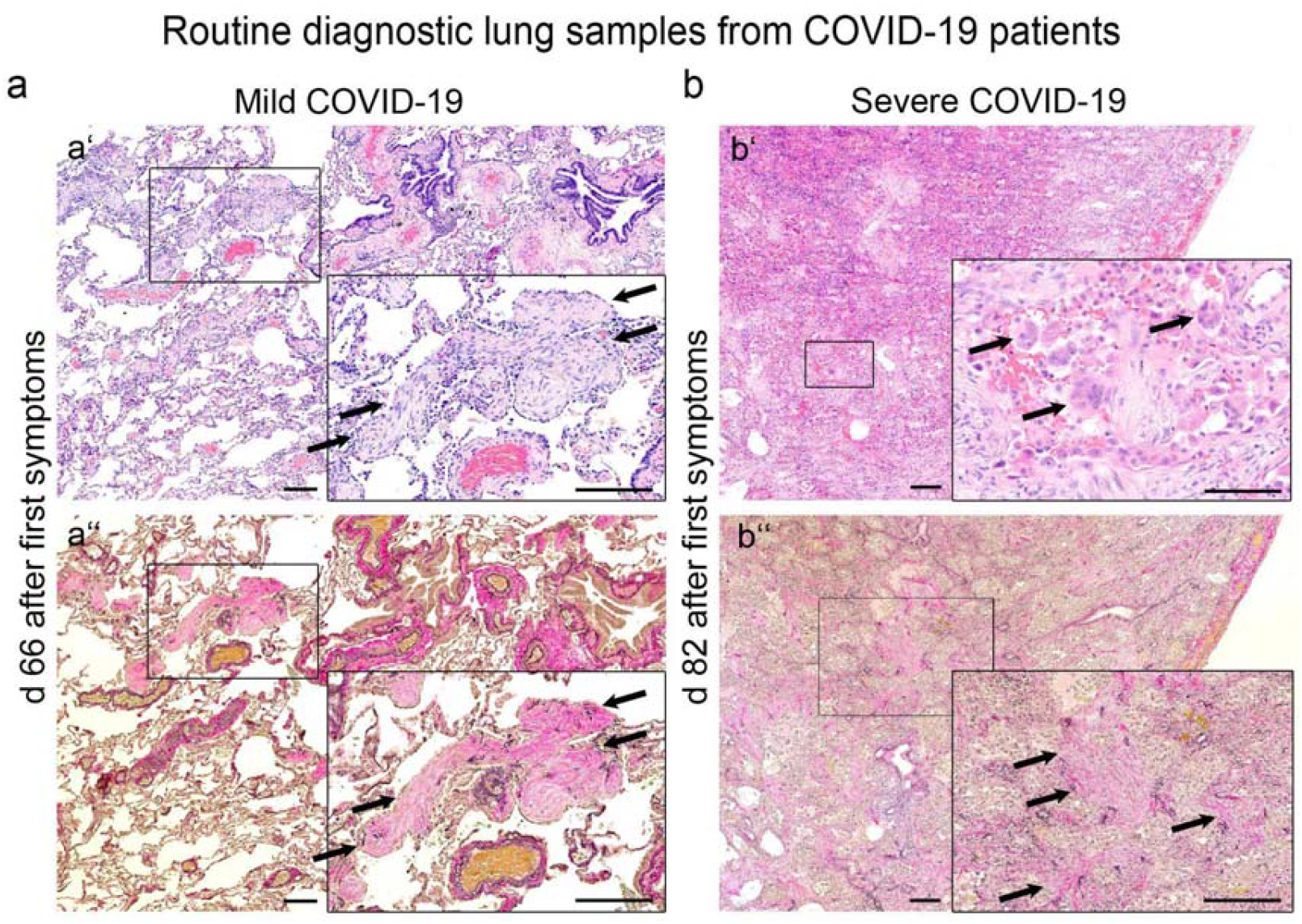
Histological findings in lungs from patients in the late phase of COVID-19. (a) Histologic images of lung tissue from a patient with lung surgery for pulmonary metastases who had recovered from mild COVID-19 (i.e., without the need for mechanical ventilation), 66 days after initial symptoms. Note the circumscribed fibrotic areas surrounded by thin alveolar septa with open alveolar spaces (arrows, a’ hematoxylin-eosin, a’’ elastica van Gieson stain, scale bar = 200 µm). (b) Histological images of lung tissue from a patient with severe COVID-19 (i.e., impaired oxygenation and subsequent failure of mechanical ventilation with the need for extracorporeal membrane oxygenation (ECMO)) with bacterial superinfection. The patient underwent surgery on day 82 after initial symptoms for histologic evaluation of fibrotic changes in the lungs. Note disappeared macrophage-filled alveolar spaces and multinucleated giant cells (b’ arrows, hematoxylin-eosin, scale bar = 250 µm, insert: scale bar = 100 µm) and diffusely fibrotic alveolar septa (b’’) arrows, Elastica-van Gieson stain, scale bar = 250 µm, insert: scale bar = 250 µm).

### SARS-CoV-2 detection in the Pandemic Peak Cohort

SARS-CoV-2 RNA was detected in two samples from the pandemic peak period in a previously unrecognized COVID-19 patient (Figure 2a). The positive samples were bilateral tonsillectomy specimens from a young female patient with clinical symptoms of tonsillitis and peritonsillar abscess formation (Figure 2b’’, 2c’’ and 2d). She remained without symptoms typical for COVID-19. The remaining N = 221 specimens from the head and neck, colon, lung, and kidney during the pandemic peak were negative (Supplementary Table 2,Supplementary Table 3).

### SARS-CoV-2 detection in the Pre-Pandemic Cohort showed no positive cases

None of the samples in the pre-pandemic cohort were positive for SARS-CoV-2 RNA (Figure 2a).

## Discussion

This study established and validated SARS-CoV-2 RNA detection methods in non-autopsy FFPE tissues, showing its utility as a prospective and retrospective screening method and a tool to investigate organ tropism and effects of SARS-CoV-2. We used commercially available kits, that enable broad applicability, and validated the analytical efficiency and limit of detection, as suggested previously (Vogels, et al., 2020). We established a dual-target assay using the E gene assay (TaqMan) as first-line screening and the RdRp gene assay (TaqMan) as a confirmatory test, both assays in a singleplex approach, are in line with a previous study (Corman, et al., 2020). A dual-target assay is included in most commercially available molecular diagnostic kits for SARS-CoV-2 RNA detection to increase sensitivity and specificity, but single-target assays are also available (Afzal, 2020). Our results show typical analytical RT-PCR efficiencies for the target E and RdRp gene assays, and the detection limit of the RdRp gene was higher than that of the E gene. When comparing the Ct values of the E and RdRp gene assays of the SARS-CoV-2 RNA standard dilution with each other, we found that the Ct values at the same viral copy number for the RdRp gene were generally higher than the Ct values of the E gene, indicating lower virus detection rates by the RdRp gene assay. A possible reason for this observation could be the detection of genomic and subgenomic mRNA, since the RdRp gene is present only in genomic mRNA (ORF1b) and the E gene is present in the genomic and also in subgenomic mRNA (Alexandersen, et al., 2020, Kim, et al., 2020). Therefore, we used the E gene standard to determine the viral load in samples. Previous studies attributed the lower virus detection to mismatch of the RdRp reverse primer and described the assay as not reliable for samples with <1000 copy numbers μL^-1^ (Corman, et al., 2020). In contrast, we found accurate viral detection up to a viral copy number of 3 μL^-1^ using the RdRp gene assay. However, in the clinically confirmed COVID-19 patient cohort, we found four positive samples using the detection of the E gene, two with a high viral load (>80 viral copies μL^-1^) and two with a low viral load (<5 viral copies μL^-1^), whereas both samples with low viral load were undetectable using the RdRp gene assay. This suggests that detection of the RdRp gene in RNA isolated from FFPE samples is less efficient compared to the SARS-CoV-2 RNA standard, but the RdRp gene can be detected up to at least 80 viral copies μL^-1^. Taken together, our established SARS-CoV-2 RNA detection method provides a reliable screening method for RNA isolated from FFPE specimens.

The virus detection method can be used to analyze viral spread in different organs and disease mechanisms by correlating pathological and molecular findings with virus presence in specimens from previously confirmed COVID-19 patients. Previous studies in autopsies from deceased COVID-19 patients suggested that a low to very low viral load can be detected in extrapulmonary organs (Best Rocha, et al., 2020, Menter, et al., 2020, Polak, et al., 2020, Puelles, et al., 2020, Sekulic, et al., 2020, Wichmann, et al., 2020). However, these are all severe and fatal cases, and it remained unclear whether such viral spread is also found in less severe and non-fatal cases. For this, analyses of pathology specimens might likely be the only available approach. In two out of 10 patients (20%) with a mild disease course not requiring mechanical ventilation, we detected SARS-CoV-2 RNA in two gastric biopsies at a very early stage of infection (one day before and one day after the first positive swab, respectively). To our knowledge, detection of viral RNA in gastric tissues has previously not been reported. This is likely because mostly postmortem tissues were analyzed, which are usually subject to strong autolytic changes in the stomach. We detected SARS-CoV-2 RNA in two out of 12 patients (17%) with a severe disease course requiring mechanical ventilation. We also identified a SARS-CoV-2 RNA-positive FFPE sample from a pleural effusion cytospin preparation of a severely ill COVID-19 patient in the early phase of infection (<21 days after first symptoms, patient 12, Table 4). Because the sample contained predominantly macrophages, this suggests the presence of the virus in these cells. We confirmed and visualized this finding with direct detection of viral RNA by FISH in these cells, in agreement with previous reports showing SARS-CoV-2 RNA in pleural effusion fluid (Lescure, et al., 2020, Mei, et al., 2020). Finally, we found a positive colonic resection specimen, interestingly in a late-stage 57 days after the first positive test, confirming previous reports of positive RNA detection in the colon and wastewaters (Lamers, et al., 2020, Lodder and de Roda Husman, 2020, Remmelink, et al., 2020). We found no other SARS-CoV-2 RNA-positive cases in the remaining 30 samples from 18 confirmed COVID-19 patients.

Of the ten PCR-negative samples from COVID-19 patients with severe symptoms, one patient was in an asymptomatic early phase of the disease (one day before the first symptoms), and nine were in a late phase of the disease, i.e. >21 days after the first symptoms. In the late phase of the disease, the virus was likely already eliminated from the organism by the immune system, and therefore was not detectable in the colon or the lungs. The negative SARS-CoV-2 RT-PCR of the lung samples, some of which showed progressive circumscribed or organized diffuse alveolar damage (Figure 3), suggests ongoing tissue damage in the absence of the virus and thereby might help to differentiate direct effects from sequelae of COVID-19. The data also confirm the concept of fibrotic pulmonary late effects of COVID-19 (Polak, et al., 2020).

We investigated the feasibility of using pathology specimens to screen for SARS-CoV-2. We did this because a) even during the first pandemic peak in our region, SARS-CoV-2 testing was only performed when patients were symptomatic, whereas some infections are known to be mild or even asymptomatic, b) false-negative results, especially depending on the stage of the disease, are not uncommon, c) no additional patient intervention is required to perform this test, d) in case of a positive result, improved tracing of infection would be possible, e) in case of a positive result, additional and more complex molecular analyses on these tissues would be possible, e.g. cell-specific virus detection using FISH. For this analysis, we focused on the period of the first local pandemic peak and on tissues that were most frequently positive for SARS-CoV-2 RNA in previous studies. Interestingly, among the n = 223 samples analyzed, we found one previously unrecognized positive patient who underwent bilateral tonsillectomy. In addition to identifying new COVID-19 patients, this might also help to improve occupational hazard assessment for staff involved in specimen acquisition, e.g. in surgery, and processing, e.g. in pathology. The analysis of SARS-CoV-2 RNA can provide information on potential exposure to such unrecognized infectious specimens, which is particularly important in perioperative diagnostics, biobanking, or cytology, where fresh unfixed specimens are processed directly.

Finally, this method also allows retrospective studies to be performed on archived FFPE material. A previous study reported a retrospective positive SARS-CoV-2 RNA detection in a respiratory specimen from December 2019 in France, well before the onset of the pandemic in Europe (Deslandes, et al., 2020). In our cohort focusing on the most affected lung tissues, we did not find a single positive case between December 2019 and February 2020.

## Limitations

The study has several limitations. These include the single-center design and the relatively small sample size, especially for samples from COVID-19 confirmed patients. Despite this, our screening could identify a previously unrecognized case of COVID-19 and also indicated no viral spread in mild COVID-19 cases. We believe that future larger and multicenter studies could further address these limitations, potentially in a similar approach to COVID-19 autopsies, for which we recently established the first national registry aimed at collecting data from all COVID-19 autopsies in Germany to enable large-scale analyses (www.DeRegCOVID.ukaachen.de) (von Stillfried, et al., 2020).

In COVID-19 positive patients, we did not have information on the date of first symptoms in six of the 22 cases, limiting the interpretation of these data. Another limitation is the reduced number of specimens sent for histopathologic analysis during the lockdown. Starting in calendar week 13 in 2020, all hospitals were asked to postpone all elective procedures to keep ICUs free for COVID-19 patients and minimize the risk of transmission to patients within the hospital. This was reflected in a 12% reduction in pathology specimens received at our center compared with the same time in previous years (data not shown), which has also been reported previously (Stathonikos, et al., 2020). Besides, the surgical staff was reduced to minimize potential exposure to infection, and many patients were also reluctant to seek medical attention during this time (Dinmohamed, et al., 2020). Even though our center was located in one of the hardest-hit early hotspots in Germany, the number of cases was still very low compared with pandemic activity in some other countries. Therefore, our results cannot be generalized to other centers, countries, or pandemic situations. Another limitation includes the use of FFPE material itself. Formalin is known to result in RNA degradation, which might reduce the sensitivity. However, in positive cases, SARS-CoV-2 RNA was detectable even at higher dilutions, when RNA concentration was not detectable or the RNA was strongly degraded, e.g. in autopsy cases.

Finally, our assay approach was not quantitative in terms of viral RNA copy numbers. Accurate quantification is probably not well possible for these samples given the variability in the preanalytical phase and thus potential virus and RNA degradation, i.e., unclear and variable time between surgery/sample collection and formalin fixation and unclear and variable duration of formalin fixation itself. However, from a diagnostic point of view, a dichotomous positive/negative result seems to be sufficient for most applications.

## Experimental procedures

### Histology samples & cohort description

We routinely analyzed diagnostic FFPE samples archived at the Institute of Pathology, University Hospital Aachen. The diagnostic process was completed for all samples. The study was given ethical approval by the ethics committee at the medical faculty of RWTH Aachen University (EK 304/20, EK 119/20, and EK 092/20). Details of the total number of identified samples, selection and exclusion criteria, topography of the samples, and final numbers of selected samples included in the study are shown in Supplementary Figure 1 and outlined in the Supplementary Methods in more detail.

### RNA isolation from FFPE specimens and SARS-CoV-2 RNA detection

We extracted RNA from FFPE tissue using a Maxwell® 16 LEV RNA FFPE Purification Kit (Promega GmbH, Walldorf, Germany) on the Maxwell® 16 IVD instrument (Promega GmbH) or with the ReliaPrep™ FFPE Total RNA Miniprep System (Promega GmbH) according to the manufacturer’s instructions. The workflow for sample preparation is shown in Supplementary Figure 2. We stored the RNA samples at -80°C until further processing.

We examined two different kits for RT-PCR analysis. We used the RealStar® SARS-CoV-2 RT-PCR Kit 1.0 (altona Diagnostics GmbH, Hamburg, Germany) for qualitative multiplex detection of the S gene (encoding spike glycoprotein) of SARS-CoV-2 and the E gene (encoding envelope protein) of lineage B beta-coronavirus (B-βCoV) using probes labeled with fluorescent reporters and quencher dyes. Internal control was included in the master mix as a PCR inhibition control. RT-PCR was performed according to the manufacturer’s instructions.(Lohse, et al., 2020, Visseaux, et al., 2020) In addition, we used the TaqMan™ Fast 1-Step Master Mix (Thermo Fisher Scientific GmbH, Dreieich, Germany) for the qualitative detection of the E gene, the RdRp gene (encoding the RNA-dependent RNA polymerase gene) and the N gene (nucleocapsid protein gene) by primer and probe sets labeled with fluorescent reporters and Quencher dyes. We used TaqMan® Exogenous Internal Positive Control reagents (Thermo Fisher Scientific GmbH, Dreieich, Germany) as internal PCR controls. We tested for TaqMan™ singleplex and multiplex assays. RT-PCR was performed according to a previous publication (Remmelink, et al., 2020). The primer and probe sequences used and the corresponding concentrations for RT-PCR are listed in Supplementary Table 5.

### Evaluation of SARS-CoV-2 RNA detection by SARS-CoV-2 RNA control

We evaluated the two RT-PCR kits and the different primer and probe sets with the Amplirun® SARS-CoV-2 RNA control (bestbion dx GmbH, Cologne, Germany) provided with 13000 viral RNA copies μL^-1^. We prepared a 2-fold dilution series with 12 concentrations. We tested the multiplex approach of the RealStar® kit, the singleplex approach of the TaqMan™ kit with each of the E, RdRp, and N genes, and the multiplex approach of the TaqMan™ kit with the E and RdRp genes in combination. To further evaluate the methods, we used diluted RNA isolated from FFPE tissue from clinically confirmed COVID-19 positive autopsies (lung, trachea).

### Fluorescence *In Situ* Hybridization (FISH)

We deparaffinized freshly cut 1-μm-thick paraffin sections from the RT-PCR-positive cytology sample and a clinically confirmed COVID-19-positive autopsy lung sample, followed by dehydration with 100% ethanol. We performed FISH on the sections using the RNAscope® Multiplex Fluorescent Reagent Kit v2 assay (Advanced Cell Diagnostics, Inc., Hayward, CA, USA) according to the manufacturer’s instructions.

### Statistical and Data Analysis

Using a SARS-CoV-2 RNA standard, we created a 2-fold dilution series to determine a standard curve for each RT-PCR assay by linear regression of the measured Ct values as a function of log viral copy number (Supplementary Figure 3). We determined the detection limit of each RT-PCR assay based on the y-intercept, making the smallest unit of a copy number detectable (10^0^ = 1) (Vogels, et al., 2020). Using the slope, we calculated the RT-PCR efficiency E according to the equation E=100x(−1+10^−1/slope^) (Vogels, et al., 2020).

## Data Availability

The datasets generated during and/or analysed during the current study are not publicly available due to protect study participant privacy, but are available from the corresponding author on reasonable request.

## Author contributions

SVS, ED, and PB designed and oversaw the study. SVS, SV, and PB wrote the initial draft of the manuscript. SVS, RDB, and SV assembled the cohort and collected material. SV carried out RNA extraction and RT-PCR experiments. SVS and RDB carried out histological analyses. PB had the conceptual idea, outlined, and financed the study. All authors read and approved the final version of the article.

## Acknowledgments

The excellent technical help of Inge Losen and Cherelle Timm is gratefully acknowledged.

## Funding

This work was supported by the German Registry of COVID-19 Autopsies (www.DeRegCOVID.ukaachen.de), funded by the Federal Ministry of Health (ZMVI1-2520COR201), by the Federal Ministry of Education and Research within the framework of the network of university medicine (DEFEAT PANDEMIcs, 01KX2021 and STOP-FSGS-01GM1901A), the German Research Foundation (DFG; SFB/TRR57, SFB/TRR219, BO3755/3-1, BO3755/9-1, and BO3755/13-1), and the RWTH START-Program (125/17).

## Disclosure

The authors declare that there is nothing to disclose.

## Notes

### Competing Interest Statement

The authors have declared no competing interest.

### Author Declarations

The study was given ethical approval by the ethics committee at the medical faculty of RWTH Aachen University (EK 304/20, EK 119/20, and EK 092/20).

