## Supplemental Methods and Tables for "SARS-CoV-2 RNA screening in routine pathology specimens"

### Supplementary Material

#### Supplementary Methods

#### COVID-19 Patients Cohort.

#### Using ICD-10 code U07.1 (COVID-19, virus identified) or clinical information on the histology request form, we identified all specimens treated at our center between March 16, 2020, and July 3, 2020, and between September 1, 2020, and October 10, 2020. The selection workflow is shown in Supplementary Figure 1. Patient characteristics are shown in Table 3 and Table 4. Sample topography is shown in Supplementary Table 1.

#### Pandemic Peak Cohort.

#### We selected samples from the pandemic peak in our region, i.e. samples received from March 16 to 27 (calendar weeks 12 and 13). We selected the tissues most likely to be virus-positive based on previous publications on viral tropism, i.e. samples from lung and respiratory tract, head and neck region, colon, rectum, and kidney (Supplementary Figure 1).

#### Sample characteristics are shown in Supplementary Table 2 and detailed sample topography is shown in Supplementary Table 3.

#### Pre-pandemic Cohort.

Given the data suggesting that initial SARS-CoV-2 infections may have occurred several months before the currently suspected onset of the pandemics in Europe, and the fact that the lung has the highest viral load of all organs, we identified all lung samples from before the pandemic, i.e. from December 2019 to the date of identification of the first index patient in our region (February 25, 2020). For details, see Supplementary Table 4.

For all cohorts described above, all available routine stains (mainly hematoxylin-eosin) of the samples were analyzed by experienced pathologists. We selected one FFPE block per available site/tissue type in the COVID-19-positive patient cohort or one FFPE block per case in the peak pandemic cohort and the pre-pandemic cohort. The selection was based on quantity and quality of the tissue, i.e. samples with the highest number of biopsy cores or the largest and most representative tissue sample, and a minimum of artifacts related to fixation, cauterization, or crushing were selected for further analysis. If inflammatory changes or neoplasia were described in the histologic report and the appropriate sample volume was sufficient, we selected samples with inflammation or neoplasia for analysis to capture a variety of pathologic changes. We excluded specimens with insufficient sample volume, i.e. at risk of having insufficient residual tissue for archiving after two 10-µm sections for RNA extraction. We also excluded biopsy specimens with neoplastic lesions and previous multiple sections for immunohistochemical or molecular pathology analysis, as these specimens must be retained in our department during the mandatory archival period of 15 years for possible further analysis in cancer care.

**Autopsy**

For the establishment and validation of the SARS-CoV-2 RT-PCR protocol from FFPE material and as and external positive control for FISH, we have used autopsy lung tissue of a confirmed, fatal COVID-19 case showing typical histopathological findings of COVID-19 (mainly diffuse alveolar damage). The autopsy was performed in two steps according to a modified standard protocol without brain autopsy to avoid aerosol production and with fixation of the organs in formalin for one week before the further macroscopic examination and organ dissection, sampling, and subsequent microscopical analyses.

#### Supplementary Tables and Results:

##### Supplementary Table 1: Sample characteristics for COVID-19 Patients Cohort

| Sample characteristics | N patients | N samples | Male | Female | Median age | Min age | Max age | Malignancy | Samples received as: | | | |
| --- | --- | --- | --- | --- | --- | --- | --- | --- | --- | --- | --- | --- |
|  |  |  |  |  |  |  |  |  | Biopsy | Resection | Fresh frozen | Cytology |
| C08.0, C04, Head/ neck + LN | 1 | 5 | 5 | 0 | 64 | 64 | 64 | 1 | 0 | 1 | 4 | 0 |
| C16-C22 Gastrointestinal | 9 | 11 | 7 | 4 | 68 | 53 | 75 | 0 | 3 | 8 | 0 | 0 |
| C34, C37 Lung + LN & thymus | 7 | 10 | 9 | 1 | 58 | 57 | 58 | 4 | 1 | 4 | 3 | 2 |
| C42.2 Bone marrow | 1 | 1 | 1 | 0 | 28 | 28 | 28 | 0 | 0 | 1 | 0 | 0 |
| C49 Soft tissue & synovia | 2 | 2 | 0 | 2 | 68.5 | 68 | 69 | 0 | 0 | 2 | 0 | 0 |
| C56-C58 Ovary + Placenta | 2 | 4 | 0 | 4 | 33 | 33 | 35 | 3 | 0 | 2 | 1 | 1 |
| C64 Kidney | 1 | 1 | 0 | 1 | 81 | 81 | 81 | 1 | 0 | 1 | 0 | 0 |
| ∑ | 23 | 34 | 22 | 12 |  |  |  | 9 | 4 | 19 | 8 | 3 |

##### Supplementary Table 2: Sample characteristics for the Pandemic Peak Cohort.

| Patient characteristics | N | Male | Female | Median age | Min age | Max age | Malignancy | Samples received as: | | | |
| --- | --- | --- | --- | --- | --- | --- | --- | --- | --- | --- | --- |
|  |  |  |  |  |  |  |  | Biopsy | Resection | Fresh frozen | Σ |
| C00.3- C14, C31 head/neck | 92 | 51 | 41 | 60 | 4 | 91 | 4 | 0 | 79 | 13 | 92 |
| C18-C20 colon | 104 | 50 | 54 | 57 | 16 | 91 | 12 | 75 | 27 | 2 | 104 |
| C34 lung | 22 | 13 | 9 | 66 | 24 | 82 | 8 | 9 | 9 | 4 | 22 |
| C36 kidney | 5 | 4 | 1 | 73 | 22 | 76 | 3 | 0 | 5 | 0 | 5 |
| Σ | 223 | 118 | 104 | 63 | 4 | 91 | 27 | 84 | 120 | 19 |  |

##### Supplementary Table 3: Detailed topography of samples from the Pandemic Peak Cohort.

| **Topography** | **Anatomic terminology** | **N samples** |
| --- | --- | --- |
| C00.3- C6 | oral mucosa | 51 |
| C07- C08 | salivary gland | 4 |
| C09 | tonsil | 13 |
| C10 | oropharynx mucosa | 8 |
| C11 | nasopharynx | 4 |
| C14 | pharynx mucosa | 3 |
| C18 | colon | 104 |
| C31 | sinus mucosa | 8 |
| C32 | larynx mucosa | 1 |
| C34 | lung | 22 |
| C36 | kidney | 5 |
| Σ |  | 223 |

##### Supplementary Table 4: Sample characteristics for Pre-pandemic Cohort.

| **Topography** | **Anatomic terminology** | **N samples** |
| --- | --- | --- |
| C00.3- C6 | oral mucosa | 51 |
| C07- C08 | salivary gland | 4 |
| C09 | tonsil | 13 |
| C10 | oropharynx mucosa | 8 |
| C11 | nasopharynx | 4 |
| C14 | pharynx mucosa | 3 |
| C18 | colon | 104 |
| C31 | sinus mucosa | 8 |
| C32 | larynx mucosa | 1 |
| C34 | lung | 22 |
| C36 | kidney | 5 |
| Σ |  | 223 |

##### Supplementary Table 5: Primer and probe set overview used in RealStar and TaqMan RT-PCR method.

The sequences and concentrations used in the RealStar RT-PCR are not specified by altona Diagnostics. The primer and probe sets for TaqMan RT-PCR are used according to previous research.(Corman, et al., 2020)

| Kit | Gene | Primer/Probe | Sequence† | Concentration per reaction |
| --- | --- | --- | --- | --- |
| RealStar | SARS-CoV-2 S gene | S gene | not provided by company | not provided by company |
| RealStar | B-βCoV E gene | E gene | not provided by company | not provided by company |
| TaqMan | SARS-CoV-2  E gene | E_Sarbeco_R | 5' ATATTGCAGCAGTACGCACACA 3' | 0.4 µM |
|  |  | E_Sarbeco_F | 5' ACAGGTACGTTAATAGTTAATAGCGT 3' | 0.4 µM |
|  |  | E_Sarbeco_P1 | 5' FAM-ACACTAGCCATCCTTACTGCGCTTCG-BHQ-2 3' | 0.2 µM |
| TaqMan | SARS-CoV-1,  SARS-CoV-2  RdRp gene | RdRp_SARSr-F | 5' GTGARATGGTCATGTGTGGCGG 3' | 0.6 µM |
|  |  | RdRp_SARSr-R | 5' CARATGTTAAASACACTATTAGCATA 3' | 0.8 µM |
|  |  | RdRp_SARSr-P1 | 5' Cy5-CCAGGTGGWACRTCATCMGGTGATGC-BHQ-2 3' | 0.1 µM |
| TaqMan | SARS-CoV-2  N gene | N_Sarbecco_F | 5' CACATTGGCACCCGCAATC 3' | 0.6 µM |
|  |  | N_Sarbecco_R | 5' GAGGAACGAGAAGAGGCTTG 3' | 0.8 µM |
|  |  | N_Sarbeco_P | 5' Cy5-ACTTCCTCAAGGAACAACATTGCCA-BHQ-2 3' | 0.2 µM |
|  | †W ≡ A/T; R ≡ G/A; M ≡ A/C; S ≡ G/C; FAM = 6-carboxyfluorescein; Cy5 = Cyanine 5; BHQ-2 = black hole quencher. | | | |

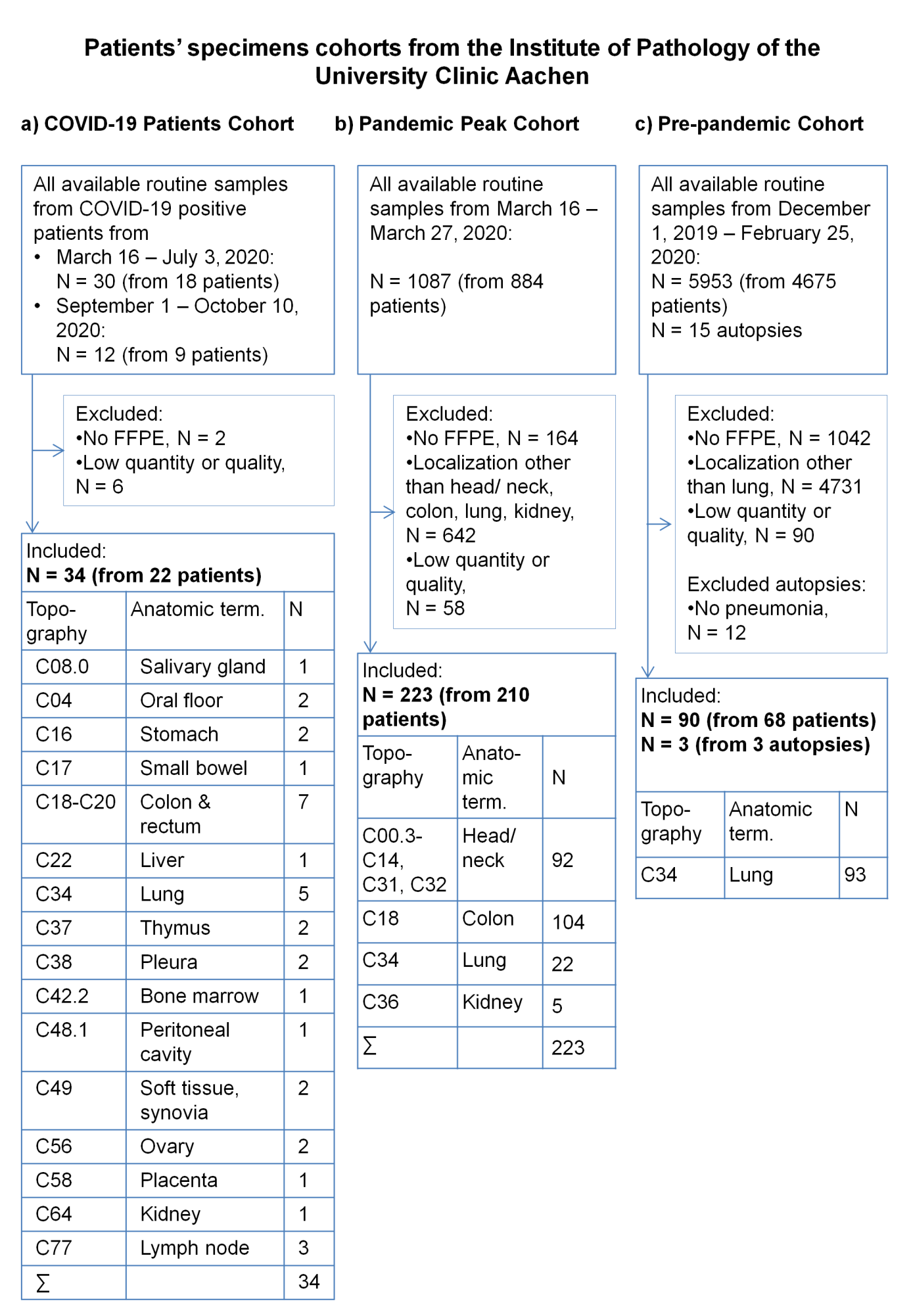

##### Supplementary Figure 1: Flow chart of sample selection of patients’ cohorts

After identifying all eligible samples, we excluded cases without the availability of FFPE material in all cohorts. Next, we excluded cases from locations other than the predefined localities in the peak pandemic cohort and the pre-pandemic cohort. Finally, we excluded FFPE samples with low tissue quantity or quality in all cohorts.

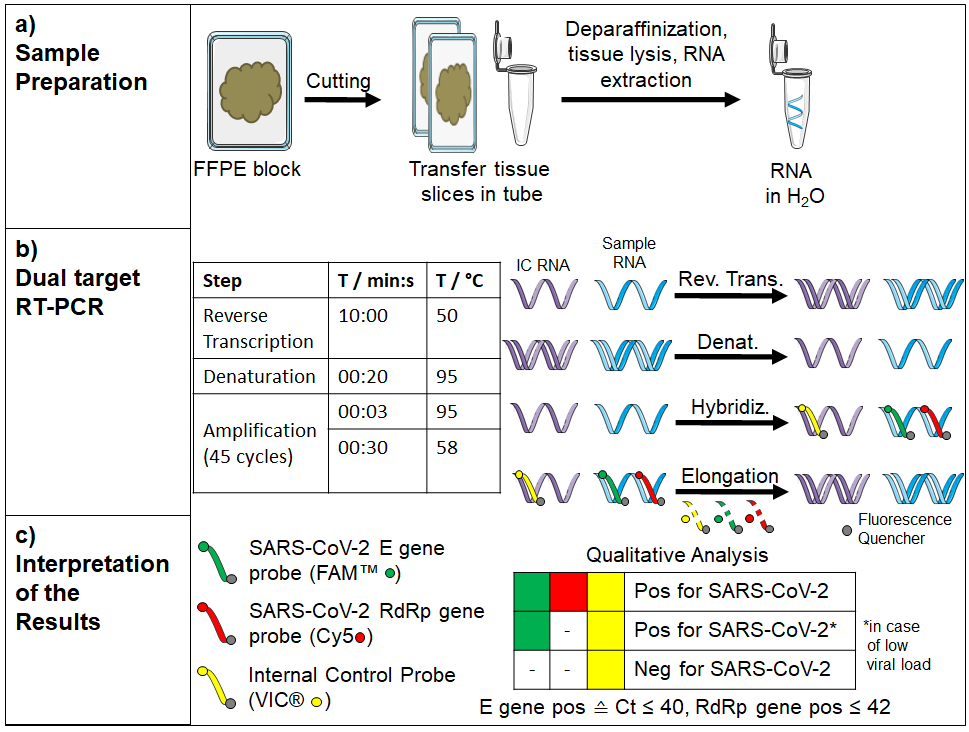

##### Supplementary Figure 2: Workflow for RNA isolation from FFPE specimens and SARS-CoV-2 RNA detection using RT-PCR.

a) RNA extraction. b) RT-PCR amplification protocol used. c) Interpretation of RT-PCR results.

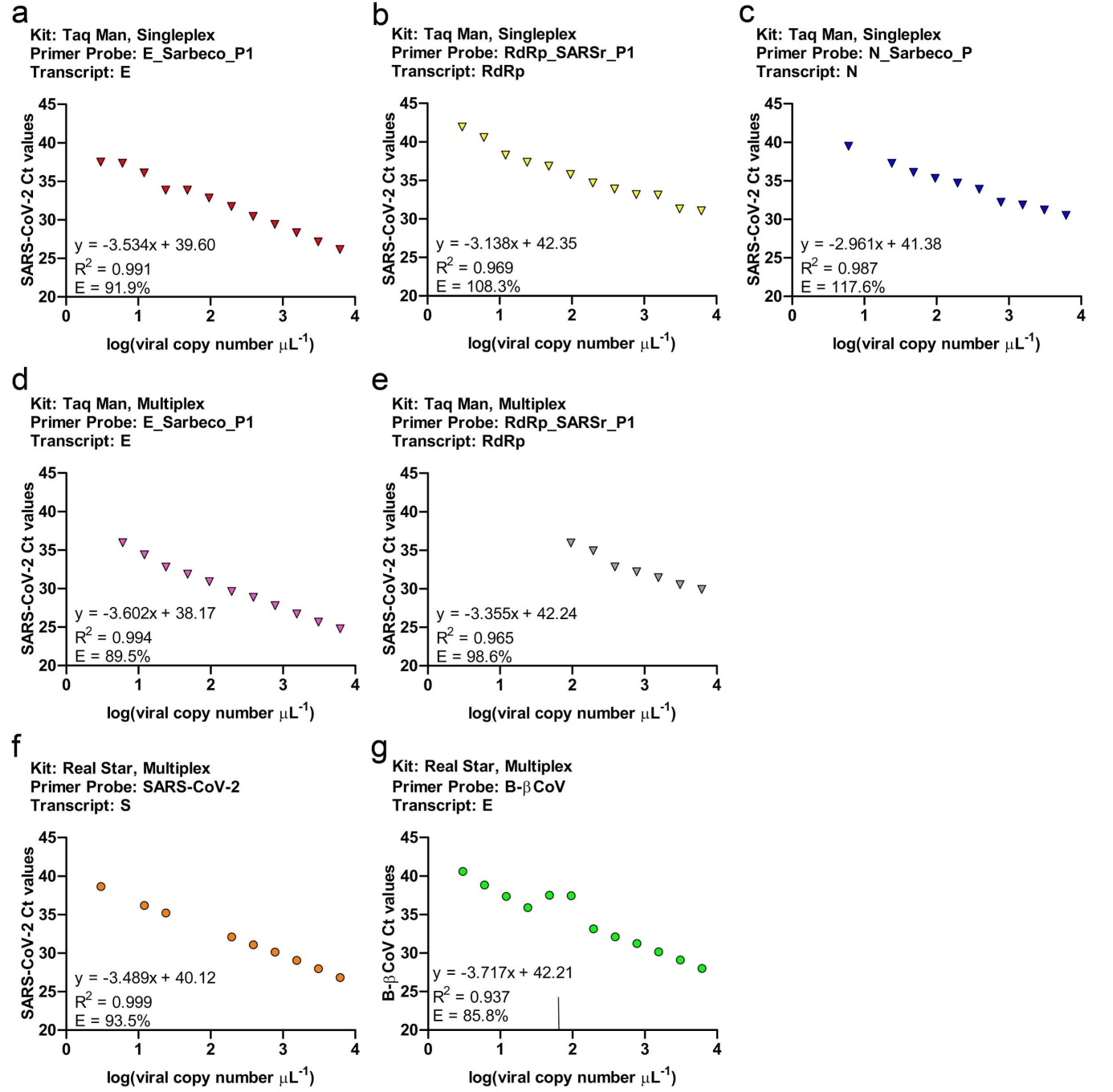

##### Supplementary Figure 3: Linear Regression and data analysis of the various RT-PCR methods.

Determination of slope, y-intercept, R^2^ and RT-PCR efficiency by 2-fold dilution series of the SARS-CoV-2 RNA standard of the approaches RealStar multiplex, TaqMan singleplex (E, RdRp and N gene) and TaqMan multiplex (E, RdRp gene).
