## Supplementary figures and images for "SARS-CoV-2 RNA screening in routine pathology specimens"

### Supplemental Figure 1

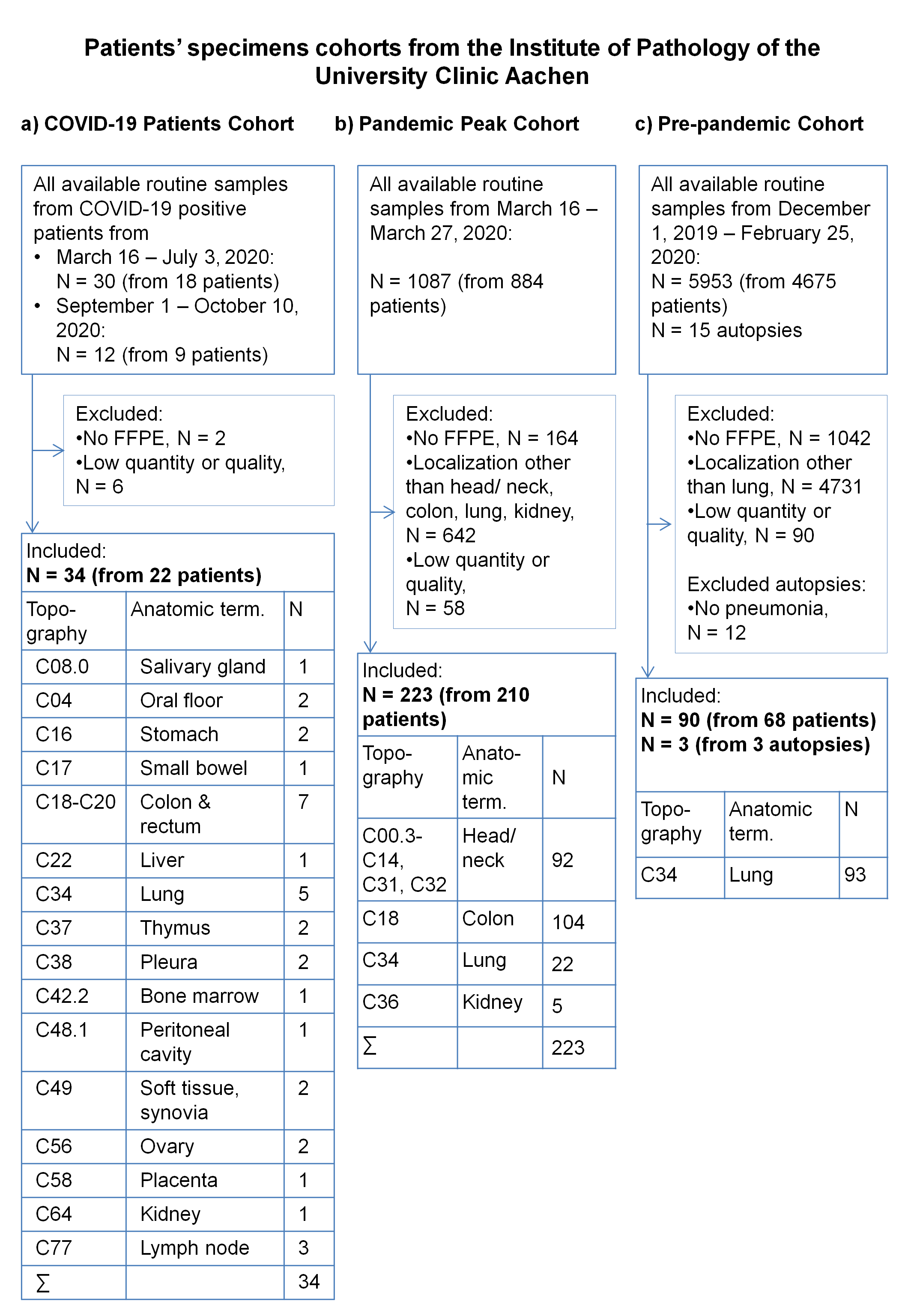

### Supplemental Figure 2

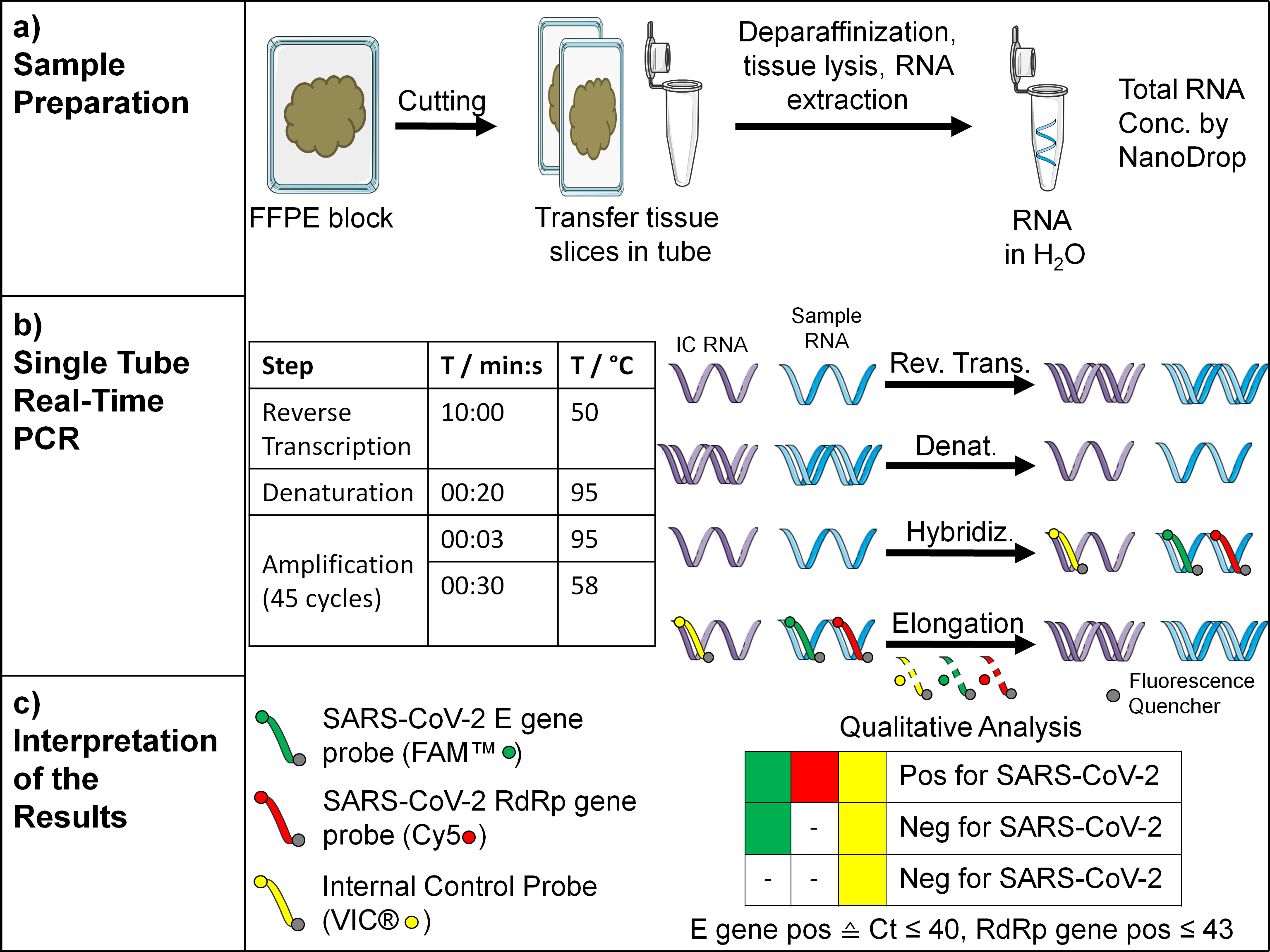

### Supplemental Figure 3

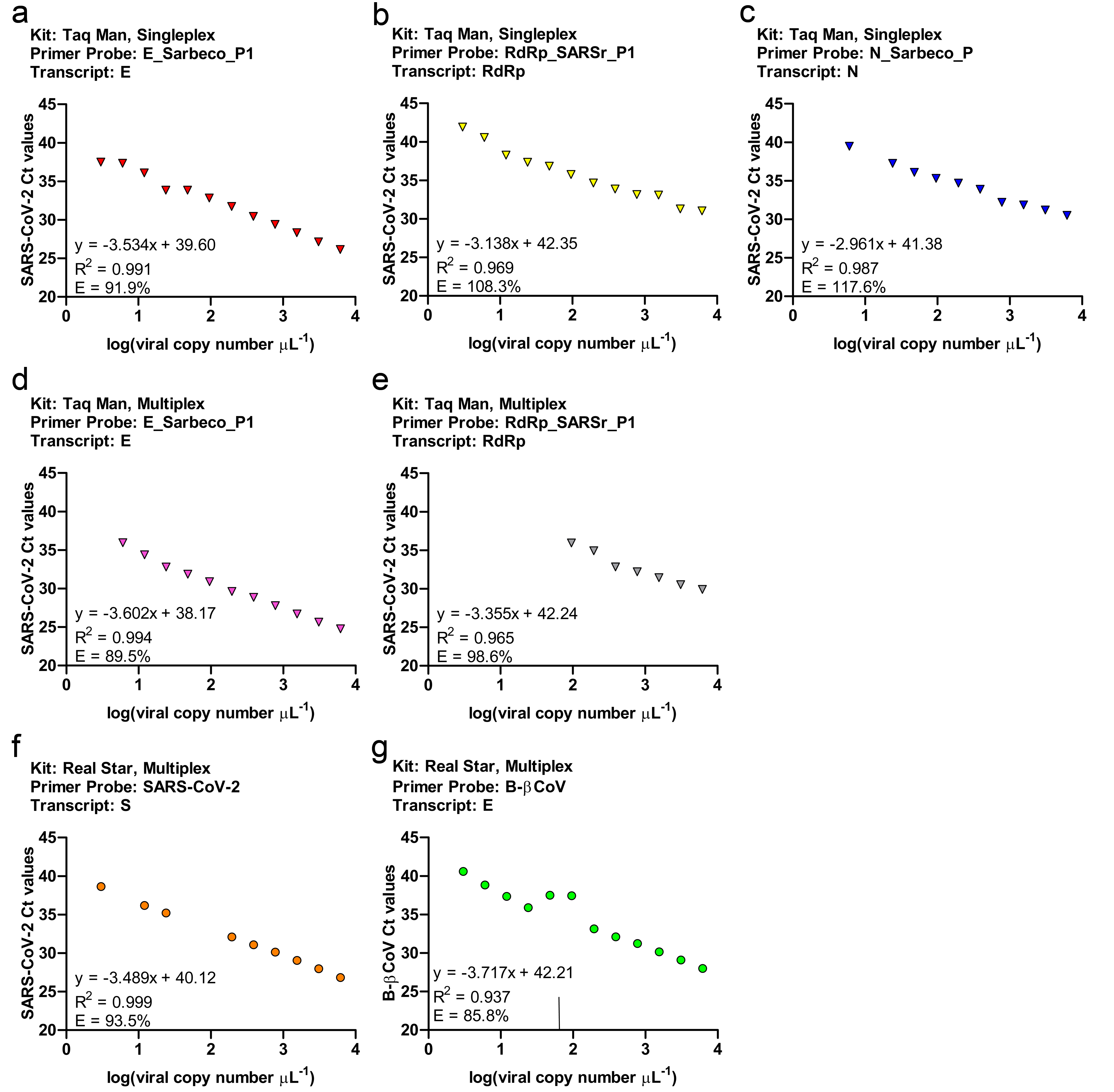
